# CLINICAL MANIFESTATIONS AND DIAGNOSIS OF CO-INFECTION OF COVID-19, TUBERCULOSIS AND OPPORTUNISTIC PULMONARY INFECTIONS IN LATE-STAGE HIV PATIENTS

**DOI:** 10.1101/2022.04.26.22274235

**Authors:** A.V. Mishina, V.Yu. Mishin, I. V. Shashenkov

## Abstract

**Objective:** The purpose of the study was to investigate the specific features of clinical manifestations and diagnosis of co-infection of COVID-19, tuberculosis and opportunistic pulmonary infections in late-stage HIV patients.

**Design:** 27 patients with co-infection of COVID-19, tuberculosis, opportunistic pulmonary infections and late-stage HIV infection with immunodeficiency without antiretroviral therapy (group 1) and 27 patients with equivalent parameters but without COVID-19 (group 2) were examined.

**Results:** The patients of the group 1 and group 2 are the persons with social maladjustment and substance addiction. All of them have concomitant viral hepatitis B/C, COPD, opportunistic pulmonary infections and similar clinical and radiological manifestations, which can only be differentiated with microbiological and molecular genetic studies. The patients with co-infection of COVID-19, tuberculosis and HIV pose a high risk of transmission of infection to healthy persons in view of non-adherence to examination and treatment.

**Conclusion:** To prevent the spread of infection among the healthy population, it is necessary to arrange in a mandatory manner an active and regular COVID-19 testing of all patients with tuberculosis/HIV co-infection, especially of late-stage HIV patients without antiretroviral therapy, in the tuberculosis care unit for HIV-infected persons at the tuberculosis dispensary.

**Setting:** There are few data on the specific features of clinical manifestations of co-infection of COVID-19, tuberculosis (TB) and opportunistic pulmonary infections (OPI) in late-stage HIV patients with immunodeficiency.

**Objective:** Study purpose is to investigate the specific features of clinical manifestations and diagnosis of co-infection of COVID-19, TB and OPI in late-stage HIV patients with immunodeficiency.

**Design:** Fifty-four (54) patients admitted for inpatient treatment at the TB Clinical Hospital N 3 named after Zakharyin were enrolled in this prospective study. The participants were assigned to two groups: group 1 (main) and group 2 (comparison group). Group 1 consisted of twenty-seven (27) aged 28-52 patients (18 men (66.7 ± 9.1%) and 9 women (33.3 ± 9.1%)) with known co-infection of COVID-19, sputum smear-positive for M. tuberculosis (MBT) pulmonary tuberculosis (PTB) and with late-stage HIV infection in progression phase without antiretroviral therapy (ART). Group 2 included 27 patients who were selected using the “copy-pair” method and were completely identical to the patients of the main group (with virtually the same age, sex, social parameters and clinical and laboratory indicators), but without diagnosis of COVID-19.

---

For etiologic diagnosis of COVID-19, SARS-CoV-2 RNA amplification with fluorescence-based real-time reverse transcription–polymerase chain reaction (RT-PCR) assay and isothermal amplification, as well as the detection of IgA, IgM, IgG antibodies to SARS-CoV-2 (including glycoprotein S receptor-binding domain) were carried out. Also, the material of the respiratory tract (nasopharyngeal and oropharyngeal swab**s**, sputum and tracheal aspirate) was studied [8].

Positive cultures for *Mycobacterium tuberculosis* (*M. tuberculosis*) (MBT) were detected in the diagnostic material of the respiratory tract (sputum, bronchoalveolar lavage, biopsy material obtained during bronchoscopy and fine-needle aspiration of the intrathoracic lymph nodes) and other organs (blood, urine, feces and punctured peripheral lymph nodes) using inoculation on solid Löwenstein–Jensen medium and automated BACTEC MGIT 960 System [9].

For etiologic diagnosis of OPI caused by *Streptococcus pneumoniae (S. pneumoniae), Haemophilus influenzae (H. influenzae), Staphylococcus aureus (S. aureus), Candida albicans (C. albicans), non-tuberculosis Mycobacteria (M. avium complex), Pneumocystis jirovecii (P. jirovecii), Herpes virus simplex (HVS) and Cytomegalovirus hominis (CMVH)* in late-stage HIV patients, virological, bacteriological, immunological methods and PCR tests were used for the study of diagnostic material of the respiratory tract obtained during bronchoalveolar lavage and bronchoscopy [10]

All patients underwent comprehensive clinical, laboratory, immunological (CD4+ T-cell enumeration by flow cytometry and viral load measurement by determination of HIV RNA copy number in the peripheral blood) and radiation examination, including computed tomography (CT), magnetic resonance imaging (MRI) and ultrasound imaging (UI) of the internal organs.

Statistical data processing was carried out using Microsoft Office Excel 2019 with the calculation of the mean of grouped data and standard errors of the mean and confidence interval (CI). P-value was evaluated using Student’s t table. Differences between the means were considered significant at p <0.05. Ethical approval was not required for the study.

## Results

In all 54 patients of the group 1 and group 2, HIV infection was the first disease; at the time of diagnosis, parenteral transmission was established in 42 (77.8 ± 5.6%) patients and sexual transmission - in 12 (22.2 ± 5, 6%) patients. All of them were registered at the AIDS centers, but due to social maladjustment and non-adherence to examination and treatment actually did not visit the centers, did not receive ART, did not work and did not have a family. All patients had drug abuse, alcohol and tobacco addiction. All of them had concomitant viral hepatitis B/C and chronic obstructive pulmonary disease (COPD) - 17 (31.5 ± 6.3%). Fifty-four (54) patients were diagnosed with PTB after 6-9 years when they presented in primary healthcare institutions with symptoms of acute respiratory disease; the diagnosis was confirmed during comprehensive examination at tuberculosis dispensaries (TD), where M. tuberculosis was detected in the diagnostic material of the respiratory tract by microscopy and inoculation on culture media, and the patients were diagnosed with PTB.

All patients were admitted to the TB Clinical Hospital N 3 named after Zakharyin with the referrals from the physicians of tuberculosis dispensaries. In the admission department of the Hospital, testing for SARS-CoV-2 was carried out; 27 patients (group 1) were diagnosed with COVID-19. They were isolated in a specialized observational unit. 27 patients (group 2) who were not diagnosed with COVID-19 were transferred to a specialized unit for patients with tuberculosis and HIV infection.

The social status of patients in group 1 and group 2 remained virtually unchanged; they continued to be substance-addicted and had viral hepatitis B/C, but the number of patients with COPD increased to 62.2 ± 6.1% (in 37 persons).

The distribution of patients in study groups by the number of CD4+ lymphocytes in 1 μl of blood is presented in Table 1.

**Table 1.** Distribution of patients in study groups by the number of CD4+ lymphocytes in 1 μl of blood (M±m)

| Group | Number of patients | Number of CD4+ lymphocytes in 1 µl of blood |  |  |  |
| --- | --- | --- | --- | --- | --- |
|  |  | 50-30 | 29-20 | 19-10 | <9 |
| 1 | Abs. 27<br>% 100 | 3<br>11,1±6,2 | 7<br>25,9±8,4 | 10<br>37,0±9,2 | 7<br>25,9±8,4 |
| 2 | Abs. 27<br>% 100 | 4<br>14,8±6,8 | 8<br>29,6±8,7 | 9<br>33,3±8,4 | 6<br>22,2±8,0 |

As shown in Table 1, the number of CD4+ lymphocytes in 1 μl of blood virtually did not differ in the study groups. In group 1, in 11.1% of patients, the CD4+ count was in the range of 50-30 cells/μL, in 25.9% - 29- 20, in 37.0% - 19-10 and in 25.9 % - < 9; and in group 2, respectively: in 14.8%, in 29.6%, in 33.3% And in 22.2 (p> 0.05).

The average number of CD4+ lymphocytes in 1 μL of blood was also the same representing 24.1 ± 0.64 and cells/μL of blood in group 1, and 29.7 ± 0.54 in group 2 (p>0.05). With that, the viral load in patients in both groups was >500,000 HIV RNA copies/ml of blood.

From there, in patients with late-stage HIV infection in progression phase without ART and with COVID-19/PTB co-infection, the CD4+ count in the blood was ≤50 cells/μl, the average number of CD4+ was ≤30 cell/μl and the viral load was > 500,000 HIV RNA copies/ml of blood; it indicates severe immunodeficiency and does not differ from the corresponding indicators in patients without COVID-19 that determines the uniformity of a poor immune response and the similarity in the disease generalization and clinical and radiological manifestations of tuberculosis.

In patients of the group 1 and group 2, PTB was combined with generalized form with multiple extrapulmonary specific lesions, confirmed by positive cultures for MBT in the diagnostic material of the various organs. With that, in group 1, two organs were affected in 12 patients, three - in 7 patients, four - in 2 patients and five - in 2 patients; in group 2, respectively: two organs - in 11, three - in 8, four - in 1 and five - in 2 (p> 0.05). The most frequent sites of extrapulmonary tuberculosis in the study groups were the intrathoracic lymph nodes and pleura (in all patients), the intestine and mesenteric lymph nodes (16 in group 1 and 17 in group 2) and the peripheral lymph nodes (18 and 18). CNS tuberculosis (6 and 5) and genitourinary tuberculosis (7 and 8) were frequent enough. Splenic TB (4 and 3) and bone and joint TB (4 and 5) were observed less frequent. Lesions of the thyroid gland, adrenal glands, pericardium and inner ear were found in single patients.

From there, in patients with late-stage HIV infection with immunodeficiency in progression phase without ART and with COVID-19/PTB co-infection, generalized tuberculosis manifests in pulmonary and multiple extrapulmonary lesions; these conditions are characterized by similar clinical manifestations and disease course.

Also, at microbiological and PCR examination of diagnostic material of the respiratory tract, pathogens of OPI were detected. The distribution of patients in study groups by clinical diagnoses and pathogens of opportunistic pulmonary infections is presented in Table. 2.

As shown in Table 2, the patients in both groups had additional OPI co-infection; the frequency and OPI pathogens did not differ significantly in the study groups.

**Table 2.** Distribution of patients by clinical diagnoses and frequencies of opportunistic pulmonary infections (M±m)

| Diagnosis of OPI | Pathogens of OPI | Number of patients | OPI frequency in groups |  | p |
| --- | --- | --- | --- | --- | --- |
|  |  |  | 1 | 2 |  |
| BP | *S. pneumoniae* | Abs. 27<br>% 100 | 11<br>40,7±9,4 | 7<br>25,9±8,4 | >0.05 |
|  | *H. influenzae* | Abs. 27<br>% 100 | 7<br>25,9±8,4 | 6<br>22,2±7,9 | >0.05 |
|  | *S. aureus* | Abs. 27<br>% 100 | 4<br>14,8±6,8 | 5<br>18,5±7,4 | >0.05 |
|  | Total | Abs. 27<br>% 100 | 22<br>81,5±7,4 | 18<br>66,7±8,2 | >0.05 |
| CP | *C. albicans* | Abs. 27<br>% 100 | 9<br>33,3±8,4 | 11<br>40,7±9,4 | >0.05 |
| PM | *M. avium complex* | Abs. 27<br>% 100 | 9<br>33,3±8,4 | 8<br>29,6±8,7 | >0.05 |
| PCP | *P. jirovecii* | Abs. 27<br>% 100 | 7<br>25,9±8,4 | 6<br>22,2±8,0 | >0.05 |
| VP | *HVS* | Abs. 27<br>% 100 | 8<br>29,6±8,7 | 7<br>25,9±8,4 | >0.05 |
|  | *CMVH* | Abs. 27<br>% 100 | 6<br>22,2±8,0 | 5<br>29,6±8,8 | >0.05 |
|  | Total | Abs. 27<br>% 100 | 14<br>51,8±9,6 | 12<br>44,4±9,5 | >0.05 |

Note: OPI - opportunistic pulmonary infections, BP - bacterial pneumonia, CP - Candida pneumonia, PM - pulmonary mycobacteriosis, PCP - pneumocystis pneumonia, VP - viral pneumonia.

Bacterial pneumonia caused by various pathogens was found in 81.5% of patients in group 1 and in the 66.7% - in group 2. In group 1, in 40.7% of cases, bacterial pneumonia was caused by *S. pneumoniae*, in 25.9% - by *H. influenzae* and in 14.8% - by *S. aureus*; in group 2, respectively: in 25.9%, in 22.2% and in 18.5% (p> 0.05). Candida pneumonia caused by *C. albicans* was found in group 1 in 33.3% of cases and in group 2 - in 40.7%; pulmonary mycobacteriosis caused by *M. avium complex*, respectively - in 33.3% and in 29.6%; pneumocystis pneumonia caused by *P. jirovecii*, respectively - in 25.9% and in 29.6% (p> 0.05). Viral pneumonia caused by *HVS* was found in group 1 in 29.6% of cases and in group 2 - in 25.9%; viral pneumonia caused by *CMVH*, respectively - in 22.2% and in 29.6% (p > 0.05). At that, 11 patients in group 1 and 9 patients in group 2 had a combination of two OPI, and respectively - in 4 and 5 patients - three OPI.

These OPIs provide specific features in clinical presentation and complicate the course of the disease; it is manifested not only in diseases of the respiratory system, but also in changes in other organs and systems-mucosal and skin lesions caused by *C. albicans, HVS and CMVH*, diseases of the abdominal organs caused by *M. avium complex*, meningitis and meningoencephalitis caused by *C. albicans* (in 6 patients in group 1 and 4 - in group 2), *HVS* (in 5 and in 6, relatively) and *CMVH* (in 8 and in 5, relatively) (p> 0.05).

From there, in patients with late-stage HIV infection with immunodeficiency in progression phase without ART and with COVID-19/PTB co-infection, incidence of OPI was the same as in patients without COVID- 19 that determines the similarity in clinical manifestations and makes it difficult to distinguish these conditions due to the simultaneous development of several pathologies with different clinical manifestations; it is required a comprehensive etiologic diagnosis of particular disease.

The clinical presentation of the disease in patients of the main group and the comparison group virtually did not differ and was characterized by a severe intoxication and general inflammatory changes with weight loss, adynamia, headache, myalgia, neuropathy, encephalopathy, palpitations, skin pallor, fever, chills, and also, markers of inflammation in laboratory tests specific to a septic state. This was also combined with symptoms of disorder of other organs and systems.

The clinical manifestations of inflammatory changes in the respiratory system in all patients also did not differ significantly and had the same-type complaints and symptoms: fever, weakness, cough, muco-purulent sputum, dyspnea and various abnormal lung sounds. It should be noted that in patients of the group 1, the cough was more intense, the mucopurulent sputum was blood-tinged; bronchospasm and progressive pulmonary heart disease took place; in some cases, there were skin rash, hypoxemia, loss of taste, smell and hearing, disseminated intravascular coagulation, thrombosis and thromboembolism. However, similar clinical manifestations were also observed with varying incidence in patients of group 2 (without COVID-19) that is determined by OPI to a significant extent.

From there, in patients with late-stage HIV infection with immunodeficiency in progression phase without ART and with COVID-19/PTB/OPI co-infection, the clinical presentation manifesting in intoxication, general inflammatory and respiratory symptoms, is virtually the same and nonspecific; the distinction can be detected only with the etiologic diagnosis of COVID-19 and OPI.

In patients of both groups, chest CT showed a complex of three pathological syndromes: dissemination, increased lung markings and adenopathy.

Pulmonary dissemination was represented by foci of varying sizes (from small to large) and intensity (from low to high) with a tendency to confluence and the formation of infiltrates; on this background, the bronchial lumens, mainly in the lower lobes of the lungs, destroyed lung tissue and bronchogenic seeding were visualized; in more than half of patients, exudative pleurisy or pleural empyema developed. The increased lung markings had a “reticular” pattern due to the development of interstitial pneumonia (resulting from lymphohematogenous dissemination) with areas of compaction of the lung tissue manifested as “ground glass opacity”, diffuse increased lung attenuation and the development of cystic and dystrophic changes; on this background, there is a thickening of the interlobar and visceral pleura. Adenopathy is represented by bilateral enlargement of the intrathoracic lymph nodes with infiltrative changes in the periphery.

In these cases, there was a simultaneous development of several pathologies of thoracic organs and changes specific to the late-stage HIV infection with immunodeficiency, including those associated with HIV infection itself - lymphoid interstitial pneumonia, nonspecific interstitial pneumonia, primary pulmonary hypertension and commonly occurring COPD resulting in emphysema, cystic and dystrophic changes, as well as right heart enlargement. At that, these changes can be associated with manifestations of OPI.

The area of affected part of the lungs in all patients was 80-100% and was virtually comparable in both groups; it was possible to differentiate the diseases on the results of the chest CT only with the etiologic diagnosis of COVID-19, PTB and OPI.

From there, in patients with late-stage HIV infection with immunodeficiency in progression phase without ART and with COVID-19/PTB/OPI co-infection, chest CT showed the same-type of simultaneously developed diseases as in patients without COVID-19. In case of co-infection, it is not possible to distinguish pathologies; it is required an etiologic verification of particular diseases based on the development of new criteria for radiological diagnosis of chest pathology and correlation with the broad-spectrum pathogen identification in patients with late-stage HIV infection with immunodeficiency.

Patients with late-stage HIV infection with immunodeficiency in progression phase without ART are diagnosed with COVID-19/PTB/OPI co-infection 6-9 years after determination of HIV infection and when they were diagnosed with sputum smear-positive pulmonary tuberculosis (PTB); all these patients of reproductive and productive age did not work, did not have a family, had drug abuse, alcohol and tobacco addiction, concomitant viral hepatitis B/C and COPD. In these patients, the disease is characterized by severe immunodeficiency (the CD4+ count in the blood was ≤50 cells/μl, the average number of CD4+ cells was <30 cell/μl), the presence not only pulmonary but generalized tuberculosis with extrapulmonary lesions and OPI caused by *S. pneumoniae, H. influenzae, S. aureus, C. albicans non-tuberculosis Mycobacteria (M. avium complex), P. jirovecii, HVS* and *CMVH* (as in patients without COVID-19).

The clinical presentation in all patients manifesting in intoxication, general inflammatory and respiratory symptoms is virtually the same and nonspecific. Chest CT showed dissemination, increased lung markings and adenopathy associated with a simultaneous development of several pathologies including those associated with HIV infection itself.

It is not possible to distinguish these diseases by clinical and radiological manifestations because in the patients with late-stage HIV infection with immunodeficiency and COVID-19/PTB/OPI co-infection, clinical and radiological manifestations have no specific features and are the same as in patients without COVID-19.

This is possible only with a special microbiological and molecular genetic examination of diagnostic material (taken not only from the respiratory system, but also from other organs) to identify the pathogens of COVID-19, PTB and OPI that is essential to early and adequate etiologic treatment.

## Conclusion

COVID-19/PTB/OPI co-infection in patients with late-stage HIV infection with immunodeficiency in progression phase without ART is characterized by generalization of tuberculosis and the presence of several OPIs, as in patients without COVID-19.

It is not possible to distinguish these conditions by clinical and radiological manifestations, and the patients with these diseases will not be identified for a long time. This category of patients poses a high risk of transmission of COVID-19 infection to healthy persons.

In light of the results obtained, in order to prevent exogenous infecting of healthy population with COVID-19, we recommend to arrange in a mandatory manner active and regular COVID-19 testing of all patients with tuberculosis/HIV co-infection, especially late-stage HIV patients without ART.

## Data Availability

All data produced in the present work are contained in the manuscript

## References

1. Blanco J, Ambrosioni J, Garcia F, Martínez E, Soriano A, Mallolas J, Miro JM. COVID-19 in HIV Investigators. COVID-19 in patients with HIV: clinical case series. Lancet HIV. 2020 Apr 15. pii: S2352-3018(20)30111-9.

2. Docherty AB, Harrison EM, Green CA, Hardwick HE, Pius R, Norman L, Holden KA, Read M, Dondelinger F, Carson G, Merson L, Lee J, Plotkin D, Sigfrid L, Halpin S, Jackson C, Gamble C, Horby PW, Nguyen-Van-Tam JS, Dunning J, Openshaw PJM, Baillie JK, Semple MG. Features of 16,749 hospitalised UK patients with COVID-19 using the ISARIC WHO clinical characterization protocol. [medRxiv preprint] https://doi.10.1101/2020.04.23.20076042

3. Gervasoni C, Meraviglia P, Riva A, Giacomelli A, Oreni L, Minisci D, Atzori C, Ridolfo A, Cattaneo D. Clinical features and outcomes of HIV patients with coronavirus disease 2019. Clin Infect Dis. 2020 May 14. pii: ciaa579. [Epub ahead of print] https://doi.10.1093/cid/ciaa579

4. Guo W, Ming F, Dong Yu, Zhang Q, Zhang X, Pingzheng Mo, Yong F, Liang Ke. A Survey for COVID-19 among HIV/AIDS Patients in Two Districts of Wuhan. The Lancet. D-20-02926. China-Manuscript. Draft-Manuscript https://doi.10.2139/ssrn.3550029

5. Härter G, Spinner CD, Roider J, Bickel M, Krznaric I, Grunwald S, Schabaz F, Gillor D, Postel N, Mueller MC, Müller M, Römer K, Schewe K, Hoffmann C. COVID-19 in people living with human immunodeficiency virus: a case series of 33 patients. Infection. 2020 May 11. [Epub ahead of print] https://doi.10.1007/s15010-020-01438-z

6. Karmen-Tuohy S, Carlucci PM, Zacharioudakis IM, Zervou FN, Rebick G, Klein E, Reich J, Jones S, Rahimian J. Outcomes among HIV-positive patients hospitalized with COVID-19. medRxiv. [This preprint report has not been peer-reviewed] https://www.medrxiv.org/content/10.1101/2020.05.07.20094797v1

7. Wu Q, Chen T, Zhang H. Recovery from COVID-19 in two patients with coexisted HIV infection. J Med Virol. 2020 May 13. [Epub ahead of print] https://doi.10.1002/jmv.26006

8. Vremennyye metodicheskiye rekomendatsii. Profilaktika diagnostika i lecheniye novoy koronovirusnoy infektsii (COVID-19). - Ministerstva zdravookhraneniya Rossiyskoy Federatsii. [Temporary guidelines. Prevention, diagnosis and treatment of novel coronavirus infection (COVID-19). - The Ministry of Health of the Russian Federation]. 2021. 206 p. (in Russian)

9. Federal’nyye klinicheskiye rekomendatsii po diagnostike i lecheniyu tuberkuleza u bol’nykh VICH-infektsiyey. [Federal clinical guidelines for the diagnosis and treatment of tuberculosis in patients with HIV infection]. M. - Tver, Triada, 2014. 56 p. (in Russian)

10. VICH-infektsiya u vzroslykh. rKlinicheskiye rekomendatsii, utverzhdennyye MZ RF / Natsional’naya assotsiatsiya spetsialistov po profilaktike, diagnostike i lecheniyu VICH-infektsii. [HIV infection in adults. Clinical guidelines approved by the Ministry of Health of the Russian Federation / National Association of Specialists in the Prevention, Diagnostics and Treatment of HIV Infection]. 2020. 530 p. (in Russian)

